# Carrying Bangladesh with Them: How Pre-Migration Healthcare Norms Shape Bangladeshi Immigrants’ Health-Seeking in the United States

**DOI:** 10.64898/2026.10.07.26364503

**Authors:** Aantaki Raisa, Andrew H. Vu

## Abstract

Bangladeshi immigrants are a rapidly growing U.S. immigrant population, yet little is known about how pre-migration healthcare experiences shape healthcare use after migration. This qualitative study explored how healthcare norms from Bangladesh influence Bangladeshi immigrants’ health-seeking behaviors in the U.S., the barriers and facilitators they encounter, and their perceptions of care quality. Semi-structured interviews were conducted with 29 Bangladeshi-born adults living in the U.S. from September 2020 to January 2021 and analyzed using the constant comparative method. Participants described healthcare expectations shaped by Bangladesh’s out-of-pocket, resource-limited system, including symptom-driven rather than preventive care, self-treatment, and family-based caregiving. In the U.S., they faced barriers such as insurance confusion, long waits, difficulty establishing care, coverage disruptions, financial concerns, and language challenges. While participants valued culturally and linguistically concordant care, preferences for Bangla-speaking or co-ethnic providers varied, especially when perceived quality was low. Community networks, online information, self-education, and self-advocacy helped facilitate care. Provider communication strongly shaped perceptions of U.S. healthcare: feeling heard, respected, and well-informed promoted trust, while dismissal or delayed diagnosis led to skepticism and avoidance. These findings suggest that Bangladeshi immigrants’ healthcare engagement is shaped by both structural barriers and pre-migration norms. Improving access will require attention to subgroup-specific histories, insurance navigation, language access, and the relational quality of care.

## Introduction

Global displacement and immigration have more than doubled in the first quarter of the 21^st^ century, from 150 million immigrants in 2000 to over 304 million in 2026 (International Organization for Migration, 2026). The U.S. hosts the largest number of immigrant individuals in the world, with 51.9 million immigrants living in the U.S. as of June 2025, comprising 15.4% of its population (Kramer & Passel, 2025). Compared with their U.S.-born counterparts, foreign-born immigrant Americans consistently report worse health outcomes, less health service utilization, and difficulties in accessing care (KFF, 2026). A meta-analysis of eight studies on U.S. immigrants found that language and cultural barriers hinder provider communication, limit understanding of medical instructions, and reduce trust in the system—contributing to unmet medical needs, delayed treatments, and worsened health outcomes (Díaz-Millón & Olvera-Lobo, 2025).

Several solutions to these problems have been posed, such as integrating professional translation services (Karliner et al., 2007), cultural competence training (Saha et al., 2008), and addressing economic barriers to healthcare (Ruiz et al., 2023). However, what remains unaddressed are the specific needs of different immigrant populations as *culture* and *language* needs are nuanced and variable (Díaz-Millón & Olvera-Lobo, 2025; Dutta & Basu, 2008; Dutta & Jamil, 2013). There is also variability in (1) home country health behaviors that immigrants tend to bring to their host country (Lee et al., 2021), (2) immigration journeys and subsequent adaptation to life in the U.S. (Castañeda et al., 2015), and (3) how social determinants of health apply to immigrants, with immigrant-specific barriers (Lui et al., 2022; Naseh et al., 2023; Ngondwe & Tefera, 2025).

The tendency to aggregate data from heterogenous immigrant populations further obscures our understanding of the specific healthcare needs of immigrant subgroups (Durant et al., 2025; Gordon et al., 2019), as the experiences of larger subgroups often mask the needs of the smaller subgroups (Gordon et al., 2019). This problem has been highlighted with the Asian population (Gordon et al., 2019), which represents 50 different countries in the continent of Asia, with diverse histories and regional relationships that influence intergroup identities (Britannica Editors, 2025; Owen et al., 2026). Moreover, Asians speak over 50 official languages (One World Nations Online, n.d.), and countries in Asia span the full range of major economic development, with a wide range of cultural perceptions of healthcare and healthcare delivery systems (*GDP by Country in Asia (2026) - IMF*, n.d.).

The current study addresses such limitations by investigating the U.S. healthcare experiences of an Asian immigrant subgroup, Bangladeshi immigrants, one of the fastest growing immigrant populations in the U.S. Specifically, we investigate their access and utilization of the U.S. healthcare system, barriers and facilitators of access and utilization, and perceptions of the system (Akhter & Yang, 2023; Pew Research, 2021). We answer the following research questions:

1. How do pre-migration healthcare norms and experiences from Bangladesh shape Bangladeshi immigrants’ health-seeking behaviors in the U.S.?
2. What barriers and facilitators do Bangladeshi immigrants face in accessing and utilizing U.S. healthcare services?
3. How do Bangladeshi immigrants perceive the quality of their healthcare encounters in the U.S. and how do these perceptions relate to their healthcare engagement?

As the overall goal of this study was to understand how immigrants from a different healthcare system—specifically, a low-to middle-income country (LMIC)—navigate the U.S. healthcare system, it is essential to first describe Bangladesh’s healthcare system (World Bank, 2024).

Establishing this context is also critical as immigrants carry their health-related behaviors, beliefs, and expectations from their country of origin into their new healthcare environment (Lee et al., 2021). Furthermore, this contextual information establishes the transferability of the study’s findings to other immigrant populations arriving from countries with similarly structured or resource-limited healthcare systems.

## Background

Bangladesh is an LMIC in South Asia with 177 million people, representing 2.4% of the world population (*Bangladesh Population Density vs World*, 2026). It is the 7^th^ most densely populated country, with 3,544 people per square mile, nearly 25x the average of 144 people per square mile in other countries (*Bangladesh Population Density vs World*, 2026). Bangladesh also has the 36^th^ largest economy with $2,593 in per capita GDP (Georank, 2026). The healthcare system in Bangladesh is both privately and publicly funded (*Healthcare System of Bangladesh*, 2023). The publicly funded system operates under the Ministry of Health and Family Welfare and provides primary and specialized healthcare services at low cost or free of charge (*Mission and Vision*, 2025). Private healthcare facilities—including private hospitals, clinics, and diagnostic centers—offer comparatively higher quality care but at much higher costs (Rahman, 2019). There are approximately 1,354 public hospitals and 5,926 private ones (Directorate General of Health Services, n.d.). In terms of distribution, there is a significant rural-urban disparity in healthcare access. Most hospitals and diagnostic centers—both public and private—are concentrated in urban areas, particularly in the capital, Dhaka (Directorate General of Health Services, n.d.).

The doctor-to-patient ratio in Bangladesh is critically low, with <1 doctor (0.7) for every 1,000 patients (World Bank Group, 2022). Patients often have to wait for days to see a clinician or spend hours to even get an appointment (Ansar et al., 2021; Moral, 2026; Tajmim, 2022).

Moreover, the Bangladeshi health system is heavily reliant on out-of-pocket payments rather than health insurance (Hasan et al., 2025). This financial model is unaffordable for many patients, particularly those with chronic or complex conditions (Saha et al., 2025). For rural patients who struggle to reach formal healthcare facilities, seeking care from local faith healers or alternative medicine practitioners often becomes the only viable option (Haque et al., 2018). These systemic issues make accessing timely healthcare extremely difficult for many (Tayeb, 2023).

Studies have also identified a lack of trust in clinicians among Bangladeshi patients (Nessa et al., 2025; Story et al., 2011). This distrust is often rooted in experiences of being prescribed unnecessary medications or undergoing unnecessary diagnostic tests (Adams et al., 2019). Additionally, due to overcrowded and understaffed hospitals, the quality of care and care maintenance are consistently poor (Mohiuddin, 2020). Patients with chronic diseases or compromised immune systems are particularly vulnerable to hospital-acquired infections, which can further deteriorate health (Zujbe et al., 2025). Consequently, many individuals come to associate hospitals with negative outcomes and actively avoid seeking care unless absolutely necessary (Amin et al., 2024; Andaleeb et al., 2007).

## Methods

This qualitative study employed semi-structured interviews to explore the healthcare experiences of Bangladeshi-born immigrants living in the U.S. The Institutional Review Board (IRB) of the [Institution name removed for review] reviewed and approved this study. Recruitment and data collection were conducted between September 2020 and January 2021.

### Theoretical Contextualization

The authors adopted a constructivist ontological approach (Lincoln & Guba, 2016), where reality is understood to be co-constructed through human interpretation and experience. Accordingly, we acknowledge that the data were interpreted through our subjective lenses and lived experiences.

The first author shares the immigrant background of the participants: born in Bangladesh, currently living in the U.S., and has English language proficiency. While this shared identity and experience offered cultural and contextual insight, the first author’s specific upbringing in Bangladesh and immigration journey differed from many participants. These experiential overlaps and divergences shaped both the engagement with the data and its interpretation.

The second author is a first-generation Vietnamese American from a family of refugees of the Vietnam War. Residing in a hub of Vietnamese immigrants in California, the second author has witnessed the cultural differences and healthcare struggles faced by this community firsthand and has assisted Vietnamese refugees in navigating the healthcare system, where many struggle to speak, understand, or read English.

### Participant Recruitment and Sampling

Eligible participants were adults who were born in Bangladesh, currently resided in the U.S., and could read and speak in English. A total of 29 participants were recruited using snowball sampling (Woodley & Lockard, 2016), leveraging the first author’s personal network and word-of-mouth strategies. Eligible individuals were contacted by telephone and provided with a study description. Interested individuals were scheduled for an online interview via Zoom, expected to last approximately 90 minutes. All individuals approached by telephone agreed to participate; no one refused.

The sample size was predetermined based on recommendations for qualitative studies with heterogenous populations, e.g., 21+ interviews (Hsu et al., 2026). While our sample appeared homogenous in terms of country of birth (Bangladesh) and language skills, participants exhibited heterogeneity in education, gender, geographic location, time in the U.S., and income. Hence, we recruited a larger sample (N=29) to ensure perspectives due to heterogeneity were captured in the data. The decision to preselect sample size rather than pursue traditional data saturation was driven by practical constraints: this study was part of the first author’s dissertation, and time, funding, and recruitment resources were limited.

## Data Collection

The first author developed a semi-structured interview guide to explore participants’ lived experiences navigating, conceptualizing, and utilizing the U.S. healthcare system. The guide consisted of three formal sections: (1) an ice-breaking segment where participants introduced themselves and discussed their identities as Bangladeshi-born individuals living in the U.S., followed by completion of a demographic questionnaire (see Appendix); (2) the core segment covering experiences with the U.S. healthcare system (topic 1), perceptions of cancer (topic 2), and feedback on a digital intervention to promote cancer awareness (topic 3); and (3) concluding remarks and instructions for receiving compensation. The depth of data generated across the three topics warrants separate publications: findings on the digital intervention have been published elsewhere (Raisa et al., 2024). The current study focuses exclusively on participants’ experiences with the U.S. healthcare system.

At the start of each interview, the interviewer (first author) reviewed the study’s purpose and procedures with the participant and obtained verbal informed consent prior to recording. Participants were not asked about protected health information or their immigration status. Interviews were conducted between September 2020 and January 2021. All interviews were conducted in English and recorded using Zoom’s recording feature, with a backup audio recorder used as a failsafe. After each interview, the first author wrote field notes summarizing the participant’s narrative and informing initial probing questions. Only one participant stopped the interview midway and completed it at a later time; no repeat interviews were conducted for other participants. Participants were compensated with $30 for completing the study.

## Data Processing and Analysis

Audio recordings were transcribed using a third-party and all transcripts were de-identified. Transcripts were imported into the qualitative analysis software, NVivo (Lumivero, 2024). The first author used the constant comparative method (Glaser, 1965) to code each transcript iteratively, developing a codebook as coding progressed. A second coder (second author) independently coded six randomly selected transcripts, representing 20% of the dataset (Stage et al., 2026). The first author and the second coder then met to discuss their coding and resolve disagreements through consensus (Stage et al., 2026). The first author subsequently organized codes into meaningful groups to identify patterns to answer each research question. The first two authors then discussed themes to finalize them.

## Ethical Considerations

All interview recordings and transcripts were stored in encrypted storage and de-identified to protect participant anonymity. Member checking, for participants to review their transcripts or the interpretation of findings (Birt et al., 2016) was not conducted because the IRB did not permit retention of participant records after the interviews and compensation distribution.

## Results

### Characteristics of Participants

The 29 participants interviewed resided in New York (13), California (6), Florida (5), Oregon (2), Minnesota (1), Georgia (1), and Texas (1). All but one participant were urban/suburban residents at the time of interview. Twenty-six participants were married at the time of interview. Participant demographics appear in Table 1.

**Table 1.** Participant characteristics (N=29)

| Characteristics | Category | n | % |
| --- | --- | --- | --- |
| Gender | Female | 15 | 51.7 |
|  | Male | 14 | 47.3 |
| Ethnicity | Bengali | 29 | 100 |
| Education | High school/some college | 3 | 10.3 |
|  | 4-year college degree | 8 | 27.6 |
|  | Postgraduate degree | 18 | 62.1 |
| Income | Below \$50,000 | 7 | 24.1 |
| | \$50,000 to under \$75,000 | 2 | 6.9 |
| | \$75,000 to under \$150,000 | 9 | 31.1 |
| | Over \$100,000 | 8 | 27.6 |
|  | No response | 3 | 10.3 |
| Health Insurance | Employer-sponsored | 13 | 44.8 |
|  | Direct purchase/Marketplace | 10 | 34.5 |
|  | Government insurance (Medicare/Medicaid) | 4 | 13.8 |
|  | Don't know/ No response | 2 | 6.9 |
| Latest Health Service Utilization | Within 30 days | 11 | 37.9 |
|  | Past 6 months | 7 | 24.1 |
|  | Past year | 4 | 13.8 |
|  | More than a year | 2 | 6.9 |
|  | No response | 3 | 10.3 |
| Household Size |  |  |  |
|  | 1-2 | 11 | 37.9 |
|  | 3-4 | 10 | 34.5 |
|  | 5 or more | 7 | 24.1 |
|  | No response | 1 | 3.4 |
| Religion | Muslim | 18 | 62.1 |
|  | Hindu | 6 | 20.7 |
|  | Non-religious | 4 | 13.8 |
|  | No response | 1 | 3.4 |
| Age (years) | Mean (SD) | 38.69 (9.64) |  |
|  | Median (Min, Max) | 38 (18, 64) |  |
| Time in the U.S. (years) | Mean (SD) | 13.69 (8.74) |  |
|  | Median (Min, Max) | 9 (4, 39) |  |

Regarding reasons for migrating to the U.S., 11 participants (37.9%) cited higher education, 10 (34.5%) family or marriage, and 8 (27.6%) better life or economic opportunities. Below we present themes and subthemes with exemplar quotes identified by participant gender, age group, and time in the US. To protect participant anonymity, participants’ ages at the time of the interview are provided into the following groups, rather than exact age: Gen Z (ages 18-23), Millennial (ages 24-39), Gen X (ages 40-55), and Baby Boomer (ages 56-74)

RQ 1: How do pre-migration healthcare norms and experiences from Bangladesh shape Bangladeshi immigrants’ health-seeking behaviors in the U.S?

### The Weight of Cultural Carry-Over

Participants described health beliefs, behaviors, and expectations from experiences in Bangladesh that influenced their health-related behaviors in the U.S. All descriptions of how the participants’ cultural practices or experiences as an immigrant from Bangladesh influenced their behaviors could be categorized into the following subthemes:

### Symptom-driven care as default

Participants mentioned need-based utilization of healthcare services rather than seeking annual (e.g., preventive) care. Such symptom-driven care included seeking healthcare only when someone was in pain, had an accident, or pregnancy-related visits:

“*I usually don’t do [checkups]. I always do it when I have an issue.*” (Female, Millennial, 10 years).

One of the participants explicitly mentioned how their healthcare practices in Bangladesh influenced their health service utilization in the U.S.,

“*If you grow up in a culture where you see that we don’t have that regular habit of just going for a doctor for just a check-up, if you don’t have anything wrong with you, right. And that kind of factors in when you move here*” (Male, Millennial, 7 years).

### Medication avoidance and self-treatment

Participants also mentioned either avoiding medicines altogether, preferring self-medication (e.g., over-the-counter medicine), or not completing medication prescriptions if symptoms resolved. One participant directly mentioned the skepticism stemming from experiences in Bangladesh, where patients often take pain killers for any health issues instead of going to a doctor:

“*All our life we use Paracetamol and get better…that was like a Paracetamol joke amongst all the Bengalis*” (Male, Millennial, 26 years).

Another participant mentioned medication avoidance stemming from experiences of lack of effective medications back home,

“*When I was growing up in Bangladesh, medicine never worked…it was ingrained in me…do not take medicine if you don’t need to*.” (Male, Gen X, 28 years)

### The intergenerational care chain

Participants described intergenerational caregiving as a recurring pattern in their healthcare experiences. We identified two distinct caregiving patterns: working-age adults providing care for elderly parents, and school-age children or college students providing care for parents.

Adult children frequently served as interpreters, navigators, and advocates for their parents, who often lacked English proficiency or health system knowledge to manage medical visits independently. Even with social English proficiency, some parents would prefer taking their children who grew up in the U.S. for medical communication:

“*In the beginning, I would take my daughter with me to my doctor appointments, even now, sometimes.*” (Female, Gen X, 8 years)

However, another participant described the intergenerational discomfort that can arise when accompanying parents,

“*When I went to the doctor with my mom, my mom couldn’t share about our sexual activity. She was embarrassed.*” (Female, Millennial, 28 years)

The same participant also mentioned how her caregiving responsibilities as a child came at a cost to her own education. Schools did not accommodate absences related to accompanying parents to medical appointments:

“*We have school where people come to attendance and they don’t care why you’re absent…your grade gets affected. So, it’s a gamble you have to play*.”

RQ2: What barriers and facilitators do Bangladeshi immigrants face in accessing and utilizing U.S. healthcare services?

Participants described a range of barriers and facilitators that shaped their access to and utilization of U.S. healthcare. These factors operated at multiple levels—individual, interpersonal, organizational, community, and policy—and often intersected in complex ways. A recurring finding was that the same factor could function as a barrier or facilitator depending on context, individual circumstances, and prior experiences.

### The Burden of System Navigation

Participants consistently described the U.S. healthcare system as confusing, slow, and difficult to learn. This burden was particularly acute in the early years after migration, but for some, it persisted even after decades in the U.S.

### Navigating insurance

Understanding the U.S. health insurance system was a reported barrier to utilizing healthcare services. Several participants attributed the difficulty, in part, to the absence of health insurance culture in Bangladesh:

“*In Bangladesh, there was no concept of insurance. You went to the doctor and you paid out-of-pocket. Given those two things, you brought that here, and you are perfectly fine, going for as long as possible without insurance or without visits to doctors.*” (Male, Gen X 28 years)

Post-migration, participants identified insurance-related jargon, such as “out of pocket,” “in network,” or “out of network,” as confusing and difficult to grasp. Some mentioned this as a barrier only during the first few years after migration,

“*Earlier on because immigrant when you come here, I didn’t know a lot of things about health insurance. Seeing a doctor was difficult at that time. But a few years later, it was everything all right.*” (Male, Baby Boomer, 39 years)

However, others continued to struggle despite being long-term U.S. residents,

“*I still cannot understand the whole detail of the insurance yet. I think now I do a little bit. There are so many options, which is probably good for many people. But it’s not very straightforward I would say to understand the insurance system here.*” (Female, Gen X, 8.5 years)

Those who displayed high insurance literacy during the interview did not describe difficulty understanding the insurance system but consistently acknowledged systemic failures in receiving care. For example, one participant, who came to the U.S. as an adolescent, accurately explained the differences between HMO and PPO plans.^1^ She also described a diagnostic delay that profoundly shaped her view of the system,

*“…there was the healthcare system’s fault. She [participant’s mother] had her first mammogram where they saw there is a tumor…They made her wait like eight months to come back again to check. And her tumor was really fast-growing. So, it went from stage zero to stage one, to stage two and three…*” (Female, Millennial, 9 years)

Another dimension of insurance-related difficulty was determining what was and was not covered,

“*Not all the employers they are giving you enough coverage to seek your medical assistance. They just give you the basic*.” (Male, Millennial, 18 years)

### Long waits for appointments and referrals

Another barrier mentioned was the prolonged waiting times to obtain an appointment and the long waits during appointments to see a doctor. One participant described a full day at the hospital,

“…*the appointment would be always in the morning, at 8 AM…and…I would come back after 4 PM, and hungry the whole day’cause early in the morning they would want me to go there without food…*” (Female, Gen X, 19 years)

### Difficulty being established as a new patient

Similar time-related barriers emerged when participants attempted to establish care, whether after migrating to the U.S. or after moving to a new state,

“*It took me three months to get assigned to a PCP for my first appointment. So it’s always difficult to find a doctor.*” (Female, Millennial, 8.5 years)

### State-wise variations

Participants also noted that moving from state to state created additional barriers. Insurance accepted in one state often was not accepted in another:

“*Though difficult trouble when we move from city to here. Their [spouse’s] insurance doesn’t accept by most of the doctors here, so they had to change their insurance*.” (Male, Gen X, 7 years)

### Language as a Nuanced but Persistent Barrier

Language functioned as a complex and context-dependent barrier. While all participants were English-proficient by eligibility criteria, communicating health issues in medical settings introduced specific challenges.

### Loss of specificity in translation

Participants described difficulty expressing health concerns with the precision needed for accurate diagnosis despite being proficient in English,

“…*lots of thing we can’t express myself in English, lots of problem. We don’t know the proper English word. But it’s the disease, so if we miscommunicate with the wrong word or the wrong language, then we will not get the proper treatment*.” (Male, Gen X, 6 years)

### Lack of Bangla interpreters

For participants who did not live in Bangladeshi-immigrant dense enclaves like New York City, interpreter services for Bangla-speaking patients were largely unavailable. One participant contrasted the availability of Spanish interpreters with the absence of Bangla interpreter services,

“*But there are no ways that we can have a translator, like most places there are no - like for Spanish speakers they have translators*.” (Female, Gen X, 8 years)

### Interpreter discomfort

While many participants with access expressed appreciation for the availability of interpreter services, some also described discomfort speaking in front of “strangers.” For these participants, having a trusted family member was preferred over a professional interpreter,

“*…you are not comfortable with talking to a stranger, right? You already are talking to a doctor. And then if you don’t have a loved one translating for you, then it becomes a whole cultural barrier of how much are you sharing?*” (Female, Millennial, 28 years)^2^

### Preference for English in medical conversations

Contrary to the general perception that immigrants prefer native language in medical settings, several participants expressed a preference for English. This preference, however, was context-specific and explained with reasoning. One reason was that some participants knew medical terminologies only in English, not in Bangla:

“*…especially if I’m taking any medical terms. I think for medical terms, English is more convenient or comfortable with me*.” (Female, Millennial, 4 years)

Another reason was the expectation that English language applies to professional settings, while Bangla is reserved for informal, social contexts:

“…*because the way they handle this thing in a very professional manner, there I prefer English, but if it’s a doctor, who I know socially, somehow, then yeah, of course, Bangla*.” (Male, Millennial, 16 years)

A third reason was that some clinicians who claimed to speak Bangla were not truly fluent, making communication in Bangla less effective than English:

“*I would rather want them to speak in English, so they actually understand rather than, if your language capacity is really beginning…*” (Female, Millennial, 9 years)

### The Protective and Constraining Role of Community

The Bangladeshi community served as both a resource and a limitation for healthcare access. Participants used community networks for doctor referrals:

“…*I’d go to Bangladeshi community because it’s a closely knit network. And then, it’s easier to find people. So, even if when I’m in a new place, I can easily find Bangladeshi people since we’re all accumulates.*” (Female, Millennial, 7 years)

Some participants mentioned finding comfort in co-ethnic providers,

“*I live in New York City and mostly around here, everyone is around my culture…the nurses over there, sometimes they’re usually Bengali and like they speak Hindi so it’s easier for us…honestly, my mom is like friends with them, so they have open conversations*…” (Female, Gen Z, 17 years)

However, community clinics often had longer wait times and, in some cases, poor quality of care that reduced trust:

“*They [Bengali doctors] are not friendly. And if you ask them some questions, they’re not gonna explain you. If you go there…fifty people are there waiting, and when you go to a doctor, he’ll say, ‘Okay, some other people are waiting for two hours. Okay, I’ll answer it later.’ Then that later time will never come.*” (Male, Gen X, 20 years)

### The Financial Tightrope

Cost was a constant concern. The uninsured (or temporarily uninsured) faced devastating bills. Those with insurance worried about copays, deductibles, and out-of-network care. The contrast with Bangladesh’s out-of-pocket system (where you pay only when you go to a clinician) shaped how participants understood and conceptualized healthcare costs in the U.S.

### Cost shock as a formative experience

For some, a single, unexpected medical cost became a defining moment that shaped their subsequent healthcare behavior,

“*I got into a car accident…then I saw the medical bill, and it freaked me out…that was very tragic moment for me. It took me some time to pay off that bill…And I never thought about healthcare much*.” (Male, Millennial, 26 years)

### Insurance as “luck” instead of a right

Many participants framed having good insurance as a matter of luck or blessing, rather than an expectation or right:

“*I have been blessed with I guess good companies that I’ve worked with who had great health benefits, so I didn’t have to really worry about the cost*.” (Female, Millennial, 15 years)

### Out of pocket costs as deterrent to care

For others, the fear of out-of-pocket expenses discouraged them from seeking care altogether: “*In some cases, it’s very tough, but when you get sick, then if you go to the doctor you have to pay more, more money. For that reason, people are discouraged to go to the doctor, they suffer in house*.” (Male, Gen x, 6 years)

### The Vulnerability of Special Populations

Certain subgroups within the Bangladeshi community faced compounded vulnerabilities that intersected with systemic barriers. This was specifically stark for individuals who reported low income or a lack of English language proficiency.

### Elder parent isolation in healthcare

Elderly immigrants faced unique challenges in navigating the healthcare system, often exacerbated by language barriers and lack of familiarity with U.S. insurance structure. One participant described the difficulties faced by his elderly parents,

“*My parents, they’re also immigrants over here. And what I noticed…they have language barrier. So they don’t feel comfortable going to any doctors. They have a very specific preference, like…the doctor needs to understand our situation, the doctor needs to have some good experience, also the things that they are explaining, they always preferred someone who could understand their language. And finding a doctor like that who has good expertise in that certain area, they would like to talk about. That was challenging for them.*” (Female, Gen X, 6 years)

### Perlis of being low-income

Low-income immigrants faced particular challenges in navigating and obtaining government-funded healthcare,

“*If I’m legal immigrant, then when I go for insurance, then I have to submit documents also with the income because I’m not older. The elderly people they no need to income to get the insurance…But for me, I need to show the income or somebody who is take care of me to submit their documents to get insurance*.” (Male, Gen X, 6 years)

Another low-income participant on Medicaid described a traumatic childbirth experience that she attributed to the way she was treated as a Medicaid patient,

“*I gave birth to two other children and I know that it was, and I still know that why I had that difficult childbirth for the first time. It is not because it was the first time, it was because they made me hold the baby from 9 AM to 12:25 PM when the baby was born. But I think about 12:20 they said that there is a bed there finally*.” (Female, Gen X, 19 years)

### Immigration status vulnerability

Participants who initially came to the U.S. on temporary visas (e.g., visitor, student) described their fear and uncertainty about seeking care due to their immigration status,

“*Initially, I was very scared about the billing’cause my husband is low-income and my sister-in-law, she lost her husband, she also had four children who’d be living with my husband too. We were like a big family living in the same apartment, not knowing about the healthcare system at all. And I was just a visitor at that time, even though I married my husband*.” (Female, Gen X, 19 years)

Non-citizen status limited access to services even for U.S.-born children with special needs. One participant explained that her son, who has autism, received fewer services because he was not a citizen:

“*I have one…son, who is a special needs…we have a health insurance, but he has a Medicaid, the state Medicaid, and he gets more services than us. But we are still not citizen in this country…so I hear that this kind of kids, they get more services in home like ABA therapy, speech therapy, and occupational therapy, but we don’t get the services*.” (Female, Gen X, 7 years)

### Immigrant Resourcefulness as a Facilitator

Despite the numerous barriers described, participants demonstrated considerable resourcefulness in navigating the U.S. healthcare system. They employed strategies that maximized the limited resources available to them and, in some cases, offered recommendations for systemic improvements.

### Identifying preferred providers

While some immigrants mentioned leaning on their community to identify suitable clinicians, others mentioned relying on online reviews,

“*First…I check with my health insurance which medical services or doctors they are going to cover. I can Google it. After that, I pick a doctor, seeing from the simple Google review, because I don’t know any doctors personally. So I have to depend on the reviews. I read the reviews and whoever I feel comfortable with, I select them and then I go visit them*.” (Female, Millennial, 4 years)

### Conducting independent research

Participants also engaged in self-education to better understand their health conditions and navigate the system:

“*First…the self-learning on the internet, how to select primary and how to go to the specialist and what could be the cost and which insurance, how insurance covers things and what could be the co-payment and how these things work*.” (Male, Millennial, 5 years)

### Self-advocacy

Another participant mentioned asserting their needs to the clinicians until they are satisfied,

“*A lot of doctors in the U.S., or somewhere, they wouldn’t do the X-ray, or do something. So be an advocate about yourself again. Tell them, ‘No, I feel more comfortable if you do an X-ray,’ or ‘if you do an ultrasound,’ or whatever that is. So I would say just, if you’re not getting the service that you need, just advocate for yourself*.” (Female, Millennial, 9 years)

### Recommendations for systemic improvement

Beyond individual strategies, participants also offered recommendations for how the healthcare system could better serve immigrant populations. One participant suggested that new immigrants should receive informational materials upon arrival,

“*I think immigrant people when they land in this country, they should be given some kind of forms or some kind of information, they should be provided with that these are the basic things they should do. We really don’t do that. Like getting early…as soon as you get into United States, you should get a doctor, registered with a doctor. All these things are very helpful*.” (Male, Baby Boomer, 39 years)

Another suggested that government subsidies should supplement employer-sponsored insurance, perceiving that employers may not have enough motivation to provide comprehensive coverage:

“*I think there should be some types of system, even though you have your health insurance from your employer, but the better treatment, you should have some other subsidiaries from the government to have you better treatment*.” (Male, Millennial, 18 years)

RQ 3: How do Bangladeshi immigrants perceive the quality of their healthcare encounters in the U.S. and how do these perceptions relate to their healthcare engagement?

Participants’ perceptions of the quality of their healthcare encounters were shaped by interpersonal dynamics, critical incidents—positive and negative—and comparisons between the U.S. and Bangladeshi healthcare systems. These perceptions, in turn, influenced their trust in providers and their ongoing engagement with the healthcare system.

### The Centrality of Interpersonal Communication

Participants did not uniformly trust or distrust providers. Instead, trust was conditional and varied by perceived empathy and the time and attention received during clinical encounters.

### Feeling unheard and dismissed

Feeling unheard or dismissed emerged as major reason for distrust or dissatisfaction with healthcare providers and the system. One participant described her frustration when her concerns were minimized by her provider,

“*I was talking to her about my acne and then she was like, she saw it, but she was like, ‘Oh, it’s nothing. You don’t need that. You don’t need to go to—’ I was asking for advice from my dermatologist, but she was like, ‘You don’t need it. Whatever you have is normal.’ Yeah, I feel like she doesn’t listen to me or she doesn’t sympathize with me like that I found in California*.” (Female, Millennial, 15 years)

### Detailed explanation as a marker of good care

Conversely, participants valued providers who took the time to explain conditions and treatments thoroughly. One participant appreciated the guidance and resources she received during her pregnancy,

“…*my doctor will guide me all through the pregnancy what’s happening in my body…They will also give me the handouts, like read this one and you will know by yourself. They’re very helpful*.” (Female, Millennial, 10 years)

### Time spent with provider as a measure of respect

The amount of time a doctor spent with a patient was perceived as a measure of respect for the patient. One participant articulated,

“*We deserve at least 20 minutes, if not more. I’m sorry, but your time cannot be five minutes…I am personally open to talk about my problems, but I know a lot of people who need a little bit longer to open up. So I would say the doctors can give people a little bit longer time…*” (Female, Millennial, 9 years)

### The Transformative Power of Good (or Bad) Encounters

Critical positive or negative incidents had a lasting impact on participants’ perception of the U.S. healthcare system and their subsequent engagement with it.

### Positive encounters shaping trust

Positive encounters, especially in initial years post-migration, or as initial experience with the U.S. healthcare system, could instill deep trust and gratitude. One father, whose new-born son required brain surgery, was highly impressed by the care his son received,

“*My experience is really good, I can tell you. Like my first son, he had a complicacy…the doctor, he was so good, so good, you can’t imagine*.” (Male, Gen X, 7 years)

### Negative encounters leading to skepticism

Conversely, encounters of mistreatment led to provider avoidance and skepticism about the healthcare system. One participant, whose mother experienced negligence resulting in a late diagnosis of breast cancer, described how the incident fundamentally changed her approach to healthcare,

“*Don’t blindly trust a doctor, which I don’t anymore, after that incident*” (Female, Millennial, 9 years)

### Comparing and Contrasting Two Health Systems

Participants’ perception of the U.S. healthcare system was also influenced by how they perceived the health system in Bangladesh. These comparisons shaped both positive and negative views of the U.S. system.

### Efficiency and superiority of the U.S. system

Some participants found the U.S. healthcare system to be more efficient to that of Bangladesh, especially those who have had negative experiences with or perceptions of the healthcare system in Bangladesh,

“*Actually here, everything is systematic you know. If you go to the doctor, they treatment you properly. They don’t give you extra medicine, but we know in Bangladesh, if you go to the doctor, they give some extra medicine*.” (Male, Gen X, 5 years)

### Perception of cost shaped by comparison

Participants’ perceptions of cost of care in the U.S. were also shaped by their prior experiences with Bangladesh’s out-of-pocket system,

“*In Bangladesh, we used to go to the doctor I’d say more frequently than in here. Here we know that we have limitations from insurance…In Bangladesh, you can basically go to a medic hospital directly and get healthcare. Here is if you go to a hospital directly, I think you’d end up in the ER, and then it could cost you a lot of money*” (Male, Millennial, 16 years).

### Explanatory communication as a point of contrast

Some participants who found a “trend” of providers in Bangladesh prescribing “unnecessary” medications, appreciated the U.S. providers explaining the reasons behind prescriptions and other treatment,

“*In Bangladesh, we would be given a medicine without any context. But here, the doctor would sit with me and explain why it works, how it works, what are the pros and cons.*” (Female, Gen X, 7 years)

### Convergence of perceptions of the two systems

Some participants initially held positive views of the U.S. healthcare system in the early years post-migration, perceiving it as superior to Bangladesh’s system given the U.S.’s reputation as a more advanced country. However, after experiencing critical negative incidents, such as diagnostic delays, dismissive care, or systemic failures, their perceptions reversed. Often, they came to apply the same skepticism they once reserved for the Bangladeshi healthcare system to the U.S. healthcare system as well:

“*I thought that U.S. doctors would be so compassionate, so good. Like they always are on your side figuring it out what’s good for you. But that’s not the truth because in Bangladesh, everybody thinks like, ‘Oh, the doctors want money,’ right? Because it’s not like here. I’m not paying them the whole amount. Our insurance is paying half or most of the money…I thought the doctor would be on your side, but after this incident, I’m always like…I always tell people, ‘Be an advocate for yourself, regardless, whatever the doctor told you. Do your own research*.’” (Female, Millennial, 9 years)

## Discussion

We explored the healthcare experiences of 29 Bangladeshi-born immigrants in the U.S., focusing on how pre-migration healthcare norms shape post-migration health-seeking behavior (RQ1), the barriers and facilitators of healthcare access and utilization (RQ2), and perceptions of the U.S. healthcare system and their influence on engagement (RQ3). We found that participants brought healthcare practices from Bangladesh—particularly symptom-driven care, medication avoidance, and intergenerational care—which often conflicted with the U.S. system’s emphasis on preventive, insurance-driven, individual care (Gorham, 2025; “Individualism, Solidarity, and U.S. Health Care,” 2012; ISPOR, 2025). Major barriers in accessing and utilizing healthcare services included navigating the U.S. healthcare system, financial concerns even when insured, and a lack of linguistically and culturally aware services. Despite these challenges, participants demonstrated significant resourcefulness leveraging social networks, online resources, self-education, and advocacy. Perceptions of the U.S. healthcare system were shaped predominantly by interpersonal communication with providers, with single positive or negative encounters having a lasting impact on trust and engagement.

### Integration with Prior Work

#### Language as a nuanced barrier

Language has been consistently identified as a barrier to healthcare access and utilization among immigrant populations (Díaz-Millón & Olvera-Lobo, 2025). However, our findings extend prior work by revealing the nuances of language as a barrier, particularly for immigrants whose languages are “less visible” in the U.S. healthcare system, such as Bangla speakers (Gonzalez-Barrera et al., 2024; Turin, Rashid, Ferdous, Chowdhury, et al., 2020; Turin, Rashid, Ferdous, Naeem, et al., 2020a). Language services may be more available in cities like New York or Detroit, where larger Bangla-speaking populations may make healthcare systems more attuned to their needs (Im, 2025). Outside these enclaves, however, interpreter services remain sparse (Chen et al., 2025; Monahan et al., 2023).

Moreover, the present study revealed that language preference is more nuanced than a simple binary. While the participants could read and speak in English—consistent with the study’s eligibility criteria—the preference for Bangla-speaking providers was pronounced, particularly among those discussing the larger Bangladeshi immigrant community or parents who were not fluent in English. However, participants did not endorse co-ethnic providers indiscriminately, emphasizing a need for genuine linguistic and cultural fluency. Bangla encompasses multiple regional dialects, and distinctions between, for example, India’s West Bengali and Bangladesh’s Sylheti variations are readily perceptible to native speakers. When a provider’s fluency is superficial, whether due to limited cultural grounding, being a non-native speaker, or being a second-generation Bangladeshi with attenuated ties to the linguistic community, participants described immediately recognizing these limitations. Given that language is inextricably linked to culture, the connotations of health-related terms may not fully translate across dialects or non-fluent speakers. Thus, while rudimentary Bangla may be preferable to no language-concordant care for those who do not communicate in English, it may be insufficient for quality care. These findings extend literature on Spanish-and Chinese-speaking immigrants’ dissatisfaction with fragmented, inauthentic language services (Berger & Peled, 2022; Ma et al., 2025).

#### Long wait times as a persistent barrier

Long wait times are a barrier to healthcare for Americans, with 40% reporting waiting longer than two months to receive care (Black, 2023). While national surveys have reported lower wait times for Asian Americans compared with other minoritized racial and ethnic groups (Caraballo et al., 2022), our study participants mentioned experiencing long wait times both in getting established as a new patient and to set up appointments for a health issue. This discrepancy highlights the need for data disaggregation. Lumping together individuals from the 50 countries (Britannica Editors, 2025) of Asia often masks subgroup needs when larger groups dominate aggregated data (Bhakta, 2022). Research similarly shows that aggregated Asian American data obscures substantial subgroup differences (Adia et al., 2020; Gordon et al., 2019).

#### Navigating insurance and system complexities

Navigating the U.S. healthcare system, specifically health insurance, has been identified as a major barrier in healthcare access for Americans in general. Termed a “complex labyrinth,” health insurance coverage has been reported to be difficult to understand among over a third of insured adults, and among two-thirds of college graduates (Pestaina et al., 2025). Our findings demonstrate how this barrier is larger for immigrants who must learn the system while adapting to other post-migration challenges. Participants described difficulties understanding insurance jargon, determining what was covered, and managing consequences of coverage gaps.

Furthermore, our finding that non-citizenship status and other migrant status-related complexities lead to discriminatory access to healthcare aligns with previous findings (Fuentes et al., 2022). This bolsters the recommendations of eliminating discriminatory healthcare policies and reducing barriers to access to healthcare regardless of citizenship status and socioeconomic backgrounds (Fuentes et al., 2022).

#### Comparison with Bangladeshi immigrants in other countries

Barriers and facilitators to preventive and primary care for Bangladeshi immigrants are sparsely documented in the U.S. Studies in Canada have explored primary care access among Bangladeshi men (Turin, Rashid, Ferdous, Naeem, et al., 2020b) while research in Australia has examined preventive services among Bangladeshi and Nepalese immigrants (Anwar et al., 2026). However, we could not identify studies specifically exploring Bangladeshi Americans’ experiences in seeking preventive or primary care.

Some of our findings align with studies from Canada and Australia: language barriers, long wait times, inadequacy of culturally appropriate health services, and limited familiarity with the system were reported across contexts (Anwar et al., 2026; Turin, Rashid, Ferdous, Naeem, et al., 2020b). The Australian study also identified similar facilitators as ours, including utilization of online resources and community networks for information and support (Anwar et al., 2026).

However, unlike these studies, we found barriers navigating health insurance and access limitation due to immigration status. These differences may reflect the distinct structure of each healthcare system. The U.S. system differs from Canadian and Australian systems in its reliance on private insurance, complexity, and cost (Gunja et al., 2023). Future research could compare healthcare systems to identify systemic features that facilitate or inhibit immigrant healthcare access and utilization.

#### New Contribution to Literature

This study makes several novel contributions to the literature on Bangladeshi immigrant health. First, we identified cultural carryover at three distinct levels—systemic (symptom-driven, reactive care), individual (medication avoidance), and interpersonal (intergenerational caregiving)—demonstrating how the structural logic of Bangladesh’s out-of-pocket, prevention-deprioritized healthcare system persists decades post-migration. Second, our findings complicate assumptions about language concordance: providers speaking Bangla did not automatically signal cultural competence; similarly, participants’ proficiency in English did not automatically transfer to proficiency in communicating culturally embedded expressions of illness in English. This finding points to the importance of cultural interpretability in healthcare communication, going beyond language proficiency. Third, family members served as both facilitators and barriers: preferred over professional interpreters yet sometimes creating intergenerational discomfort around sensitive topics. Finally, participants did not uniformly prefer co-ethnic providers; some perceived them as providing lower-quality care, weighing trade-offs between cultural alignment and perceived competence. Collectively, these findings challenge simplistic notions of cultural concordance and underscore the need for healthcare systems to move beyond tokenistic language services toward genuine, multifaceted cultural competence.

#### Transferability

Findings may be transferable to immigrants who share similar characteristics, including English proficiency, insurance coverage, and migration from healthcare systems characterized by out-of-pocket payments, limited insurance infrastructure, and government-subsidized public hospitals for low-income populations. They may also extend to immigrants from collectivist cultures where health is viewed as a shared family concern rather than solely an individual responsibility (Dutta & Jamil, 2013). However, transferability should be considered cautiously because geographic, cultural, socio-historical, and religious differences may produce divergent experiences. Most participants also migrated for higher education or economic opportunity rather than through pathways associated with asylum, persecution, or the diversity visa lottery, which may further shape healthcare experiences (International Rescue Committee, 2025; U.S. General Services Administration, n.d.).

#### Limitations

This study has several limitations. First, the sample was predominantly insured, highly educated (62.1% held postgraduate degrees), and relatively high-income (58.7% earned $75,000 or more); experiences of lower-income, uninsured, or less-educated Bangladeshi immigrants may differ substantially. Second, participants were English-proficient by eligibility criteria, and those with limited English proficiency likely face additional challenges. Third, participants were predominantly urban or suburban, whereas rural Bangladeshi immigrants may experience greater isolation and limited access to co-ethnic providers.

### Implications

#### Filling a data gap

This study addresses a significant gap in the literature by providing disaggregated data on Bangladeshi immigrants, one of the fastest-growing yet least-studied immigrant populations in the U.S. Aggregating Asian American data can obscure substantial subgroup differences, masking disparities and perpetuating the Model Minority Myth—the stereotype that certain minority groups are uniformly successful, hardworking, and high-achieving (Adia et al., 2020; Bhakta, 2022; Gordon et al., 2019; Walton & Truong, 2023). Although Asian Americans and South Asians experience post-migration stressors shaped by their distinct histories and sociopolitical contexts (Castañeda et al., 2015; Samuel, 2009), Bangladeshi-specific healthcare experiences remain scarcely studied. By centering Bangladeshi immigrants’ experiences, this study helps address this gap and provides a foundation for future research.

#### Policy implications

Policies designed to accommodate more vulnerable populations have historically proven beneficial for society as a whole. For example, the Americans with Disabilities Act (ADA) improved accessibility for everyone, not just those with disabilities (Administration for Community Living, 2026). Similarly, expanding interpreter services, simplifying insurance enrollment, and ensuring coverage portability across states could improve healthcare access for immigrants while benefiting other populations navigating complex systems. Eliminating discriminatory policies related to immigrant healthcare access is also essential to advancing health equity (Young et al., 2023).

## Data Availability

All data produced in the present study are available upon reasonable request to the authors

## Footnotes

1 HMO= Health Management Organization’ PPO = Preferred Provider Organization

2 The issue of interpreters gets exacerbated in smaller, tight-knit diasporas like Bangladeshis where there is a higher chance of an interpreter knowing the patient or being socially connected by only a few degrees of separation. Hence, patients can be uncomfortable in front of an interpreter even with HIPAA protections (MacFarlane et al., 2020).

