## Appendix for "Carrying Bangladesh with Them: How Pre-Migration Healthcare Norms Shape Bangladeshi Immigrants’ Health-Seeking in the United States"

**Demographics**

1. What is your age?

- _________

2. What is your sex?

- Male
- Female

3. How do you describe your race? (Check all that apply.)

- American Indian or Alaska Native: A person having origins in any of the original peoples of North, Central, or South America, and who maintains tribal affiliations or community attachment.
- Asian: A person having origins in any of the original peoples of the Far East, Southeast Asia, or the Indian subcontinent including, for example, Cambodia, China, India, Japan, Korea, Malaysia, Pakistan, the Philippine Islands, Thailand, and Vietnam.
- Black or African American: A person having origins in any of the black racial groups of Africa.
- Native Hawaiian or Other Pacific Islander: A person having origins in any of the original peoples of Hawaii, Guam, Samoa, or other Pacific Islands.
- White: A person having origins in any of the original peoples of Europe, the Middle East, or North Africa.

4. What language is primarily spoken in your home?

- English
- Bangla
- Other: ___________________

5. What is your marital status?

- Married
- Divorced
- Separated
- Single
- Widowed

6. What is the last grade or class you completed in school?
   - Less than grade 1
   - Grades 1-8
   - High school incomplete (grades 9-11)
   - High school graduate (grade 12 or GED certificate)
   - Technical, trade or vocational school AFTER high school
   - Some college, no 4-year degree (includes associate degree)
   - College graduate (B.Sc., B.A., or other 4-year degree)
   - Post-graduate training/professional school after college (M.A., Ph.D., JD or MD)
7. Do you have health insurance?
   - Yes
   - No
8. (If yes) What kind of insurance do you have? (Mark all that apply)
   - Health insurance through your or someone else's job
   - Health insurance bought directly by you or your family
   - Medicare
   - Medicaid
   - Other, please specify: ____________________
   - None
   - I don't know
9. Which of the following income ranges best describes your total household income in 2019 before taxes from all sources?
   - Less than $15,000
   - $15,000 to under $20,000
   - $20,000 to under $30,000
   - $30,000 to under $40,000
   - $40,000 to under $50,000
   - $50,000 to under $75,000
   - $75,000 to under $100,000
   - $100,000 to under $150,000
   - $150,000 or more
   - I'd prefer not to answer.
10. How many people are in your household?
    - 1
    - 2
    - 3
    - 4
    - 5
    - 6
    - 7
    - 8
    - More than 8
11. What is your religious preference?
    - Protestant Christian (includes Baptist, Episcopal, Jehovah's Witness, Lutheran, Methodist, Presbyterian, Episcopalian, Pentecostal, Church of Christ, etc.)
    - Catholic (includes Roman Catholic)
    - Christian Orthodox (include Greek Orthodox, Russian Orthodox)
    - Jewish
    - Muslim (includes Islam/Islamic)
    - Buddhist
    - Hindu
    - Other, please specify ____________________
    - No religion
    - I would prefer not to answer.
12. Have you screened for any of the cancers below at any time in the past? (Mark all that apply)
    - Prostate (e.g., PSA)
    - Lung (e.g., low-dose CT scan)
    - Breast (e.g., mammogram)
    - Pancreas (e.g., endoscopic ultrasound)
    - Skin (e.g., examination by doctor)
    - Stomach (e.g., endoscopy)
    - Gynecological (e.g., pap smear)
    - Other, please specify: ____________________
    - To my knowledge, I have never screened for any type of cancer.
